# Evaluation of SARS-CoV2 antibody Rapid Diagnostic Test kits (RDTs) and Real Time-Polymerase Chain Reaction (Rt-PCR) for COVID-19 Diagnosis in Kaduna, Nigeria

**DOI:** 10.1101/2020.11.24.20231324

**Authors:** Oluwafemi Ige, Ayuba Sunday Buru, Tanko Zainab Lamido, Tahir Mohammed, Livingstone Dogara, Ijei Ifeoma Patience, Bello-Manga Halima, Audu Reward, Ige Samuel, Nmadu Grace

**Affiliations:** Department of Medical Microbiology and Parasitology, Faculty of Basic Clinical Sciences, College of Medicine, Kaduna State University, Kaduna, Nigeria; Genomic Research Lab, Department of Medical Microbiology and Parasitology, Faculty of Basic Clinical Sciences, College of Medicine, Kaduna State University, Kaduna, Nigeria; Department of Medical Laboratory Science, Faculty of Health Sciences, College of Medicine, Ahmadu Bello University Zaria, Nigeria; Department of Haematology and Blood Transfusion, Faculty of Basic Clinical Sciences, College of Medicine, Kaduna State University, Kaduna, Nigeria; Department of Paediatrics, Yusuf Danstoho Memorial Hospital, Tudun Wada, Kaduna; Department of Medical Microbiology and Parasitology, BDTH, Kaduna State; Department of Community Medicine, Faculty of Clinical Sciences, College of Medicine, Kaduna State University

**Keywords:** SARS-CoV2, Rapid Diagnostic Test kits (RDTs), Real Time-Polymerase Chain Reaction (Rt-PCR), Liferiver®, Genefnder®, Innovita®, Kaduna State

## Abstract

The emergence of the RNA virus SARS-CoV2, the causative agent of COVID-19 and its declaration by the World Health Organization (WHO) as a pandemic has disrupted the delicate balance in health indices globally. Its attendant immune dysregulation and pathobiology is still evolving. Currently, real time PCR is the gold standard diagnostic test, however there are several invalidated antibody-based tests available for possible community screening. With ongoing community transmission in Nigeria, neither the true burden of COVID-19 nor the performance of these kits is presently known. This study therefore, compared the performance of the SARS CoV2 antibody test and the real time Polymerase Chain Reaction (Rt-PCR) in the diagnosis of COVID-19. For the purpose of this evaluation, we used the diagnostic test kit by Innovita® Biological Technology CO., LTD China, a total of 521 venous blood samples were collected from consenting patients for the SARS COVID-19 rapid diagnostic kit and Oral and Nasopharyngeal swabs were collected and analyzed using the real time Polymerase chain reaction technique for nucleic acid detection and quantification.

## Introduction

The outbreak of the new coronavirus (SARS-CoV-2/COVID-19) infection has spread to every continent of the globe, thus, forcing us to adapt to living with it. Since the emergence of this virus, there has been a lot of challenges in trying to curtail it, leading to a halt in all economic and social activities both locally and internationally. (Nicola *et al*, 2020).

The SARS-CoV-2 virus is highly contagious and spreads easily within populations. Factors that aids the spread of the virus include; population density, environment social culture (Kaushik *et al*, 2020). The Clinical presentations of COVID-19 range from asymptomatic/mild symptoms to severe illness leading to multiple organ failure and death. Mortality from the disease is associated with advanced age, immune-compromised states, and those with comorbidities. (Guan *et al*, 2020).

Currently, there are over 5million cases worldwide with∼340,000 deaths. In Nigeria, initial cases of COVID-19 were imported from other countries, however, with the advent of community transmission, there are 61,194 confirmed COVID-19 cases, 52,303 discharged and 1119 deaths as at 17/10/2020 (NCDC 2020). At the moment, there are no effective therapeutic strategies to treat COVID-19. Several drug therapies and vaccines are in different stages of clinical trials, however, none of these trials have given conclusive results yet. (Darrel, 2020; Ryan, 2020) Prevention and control strategies employed include; physical distancing, heightened personal and respiratory hygiene as well as prompt notification of the presence of ant case defining symptoms.

Since onset of this pandemic, schools and a lot of other social activities have come to a standstill. The big question is how long will this remain? Strategies towards gradual reopening of the society have to be developed. Testing for antibodies to the SARS-CoV-2 virus and determining the level of herd immunity is a viable strategy that will aid policy makers on deciding when to re-open societies. Despite the medical, social and economic ramification of this pandemic Nigeria in general and Kaduna State in particular relies on tests kits, reagents and other accessories produced in other countries. Best practices require that any new test kit be validated before use. This will ensure that the performance of these kits is assessed and informed-decisions are taken by policy and regulatory bodies. The COVID-19 is also associated with socioeconomic problems. There is fear of contagion of the infection among the populace; the concept of social distancing is not compatible with the Nigerian and African communal structure. The lockdown policy embarked on in many states of the Nigeria Federation is not without hardship on the populace despite economic palliative measures of the government. At some point, a decision has to be made on when to re-open the economy and allow normal activities to resume, however, there is no locally relevant data on community exposure to and recovery from SARS-CoV2 in Kaduna State. This gap that will limit policy decisions such as when to ease restrictions and allow for normal activities including social, economic and educational to recommence; as well as vaccination targets for vulnerable versus general population

In response to the rising COVID-19 pandemic, several diagnostic test manufacturers have developed rapid and easy to use test kits to facilitate testing both within and outside the laboratory. Among which are test kits for the detection of antibodies in blood samples. However, the WHO recommends the validation of these test kits in appropriate populations and settings. (WHO, 2020) This is to prevent false positive or negative categorization of people which may hinder disease control efforts.

The availability of a locally validated and adapted antibody test kit will improve the ease and access to a screening tool for point of care testing in primary, secondary and tertiary health care centres thereby, strengthening the local testing capacity for COVID-19. This will generally improve the ease of diagnosis/screening of COVID-19 in resource constrained settings. The detection of asymptomatic infections using a locally validated serological test kit will also improve community surveillance capacity, which will aid in decision making on measures to control the viral spread such as when to enforce quarantine or isolation.

The ability to detect the immune response using a rapid and reliable serology test has a huge advantage in the assessment of the viral spread in the population and the level of herd immunity. This study will shed more light in understanding the immune response to the virus in both immune-competent and immune-compromised individuals. The comparative advantage of utilization of the validated test kits for COVID-19 screening in the general community cannot be overemphasized. This will help policy makers and clinicians to pick up previously undiagnosed asymptomatic infected persons who might have recovered and developed antibodies and may serve as a reservoir for convalescence plasma to be utilized for those with severe infection as a form of passive immunity, in the absence of specific drug therapies or vaccines. The screening can also help measure and determine how the society gradually ease back to normalcy and the number of patients coming for voluntary testing as well as reduction in stigma, which are in accordance with the targets of Goal 3 of the Sustainable Development Goals (SDG). This pilot study therefore, seek to assessed and compare the performance of the SARS CoV2 antibody test and Real Time Polymerase Chain Reaction (Rt-PCR) in the diagnosis of COVID-19. (Elitza *et al*., 2020)

## Materials and Methods

### Study Design

The study was conducted at 3 sites using both experimental and cross-sectional study designs. We employed an experimental study design as described by Momeni *et al* (2018) to assess the accuracy and precision of a Candidate SARS-CoV-2 test kit, an antibody-based rapid diagnostic kit for COVID 19.

### Study Setting

Infectious diseases control center (IDCC) and Hamdala Motel COVID 19 treatment center, and in the Community, Kaduna State.

For the purpose of this evaluation, the diagnostic test kit by Innovita® Biological Technology CO., LTD China, will be used on Venous whole blood samples (4.0mls) dispensed into Ethylene diamine tetra-acetic acid (EDTA) anticoagulated bottles.

Oral and Nasopharyngeal swabs will be analyzed using the real time Polymerase chain reaction technique for nucleic acid detection and quantification, following RNA extraction (with Liferiver extraction kits, Shanghai, China) using primers obtained from Genefinders Company LTD, South Korea.

### Eligibility

Patients who present for community testing and those confirmed positive for SARS-CoV2 virus by RT PCR at the Isolation centers who give consent Assuming a confidence (accuracy) of 0.95%, reliability (precision) of 0.90% and a sample size of 521, True positive value, true negative value and positive and negative predictive value of the antibody was assessed using the formula:

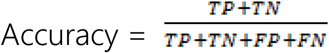

Acceptability criterion = accuracy of ≥95%

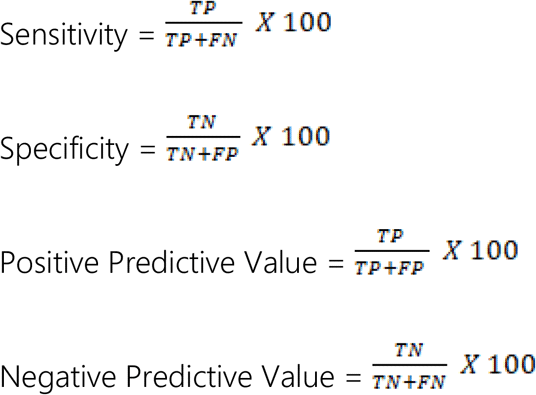

### Data Analysis

Data was analyzed using SPSS data analysis package ver. 23. Data for validation studies is presented as percentages. Qualitative daft is summarized as percentages using charts and tables. Non skewed quantitative data will be summarized as means and standard deviations (SD) while skewed data will be summarized as medians and interquartile ranges (IQR). Proportions will be compared using Z-tests for proportions. Level of statistical significance will be set at p≤0.05.

### Ethical consideration

Ethical approval from Kaduna State Ministry of Health was obtained. Privacy of participants and confidentiality of all data was ensured during the study. The study was at no cost to the participants and they were free to withdraw at any point without loss of rights or privileges.

### Data Management

All data was safely secured using alphanumeric pass-worded computers with up-to date commercial antivirus software. Each entry was backed immediately using a secure external hard drive. Hard copies were kept in a separate, safe office under lock. Only authorized individuals had access to the data.

## Results

Serum samples were tested from 521 persons 167 of whom were COVID-19 cases and 354 were healthy individuals. The mean age of the participants was 35.2± 15 years. Majority of the cases (42.5%) and healthy (39.0%) participants were less than 30 years of age. There were more male cases (60.5%) and male healthy participants (60.5%) than females.

### IgM and IgG reactivities in negative control samples

25 of sera from healthy blood donors tested IgM positive in the assay (25/354, 7.1%), while five tested IgG positive (5/354, 1.4%) (Tables 2 and 3).

**Table 1:**
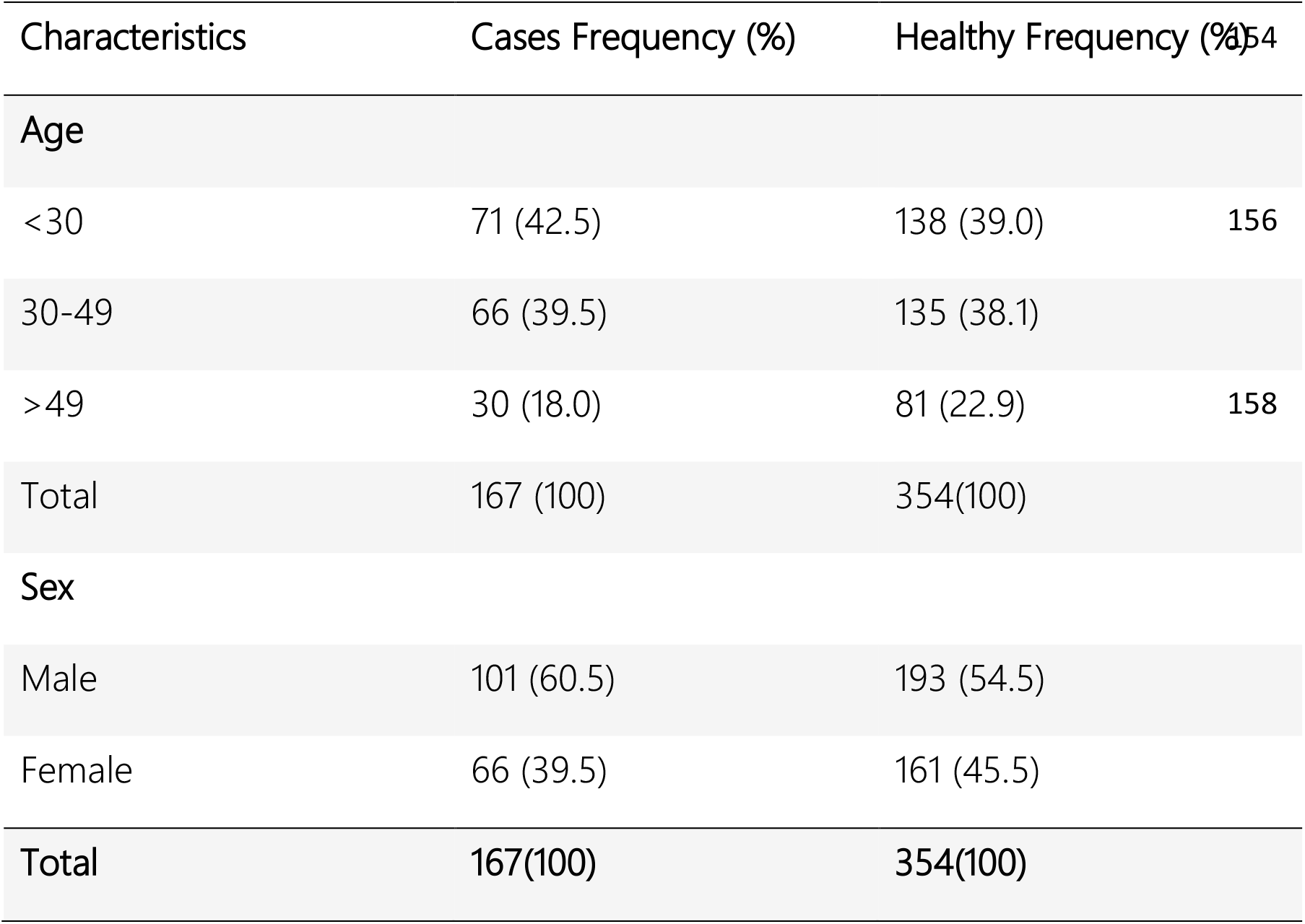
Sociodemographic characteristics of participants N=521

**Table 2.** Comparisons of IgM results for 167 PCR-positive COVID-19 cases and 354 healthy individuals

|  | Cases | Healthy | Total |
| --- | --- | --- | --- |
| IgM positive | 24 | 25 | 49 |
| IgM negative | 143 | 329 | 472 |
| Total | 167 | 354 | 521 |

**Table 3.** Comparisons of IgG results for 167 PCR-positive COVID-19 cases and 354 healthy individuals.

|  | Cases | Healthy | Total |
| --- | --- | --- | --- |
| IgG positive | 26 | 5 | 31 |
| IgG negative | 141 | 349 | 490 |
| Total | 167 | 354 | 521 |

**Table 4.**
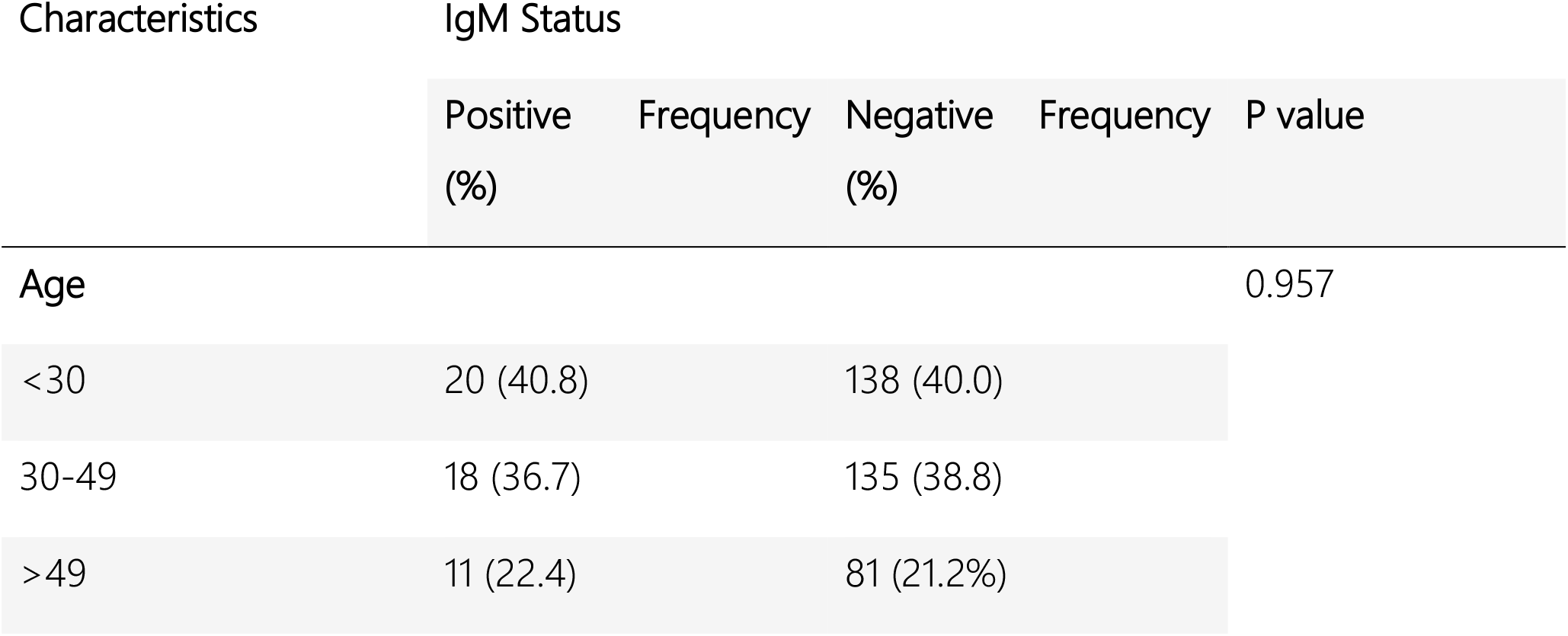
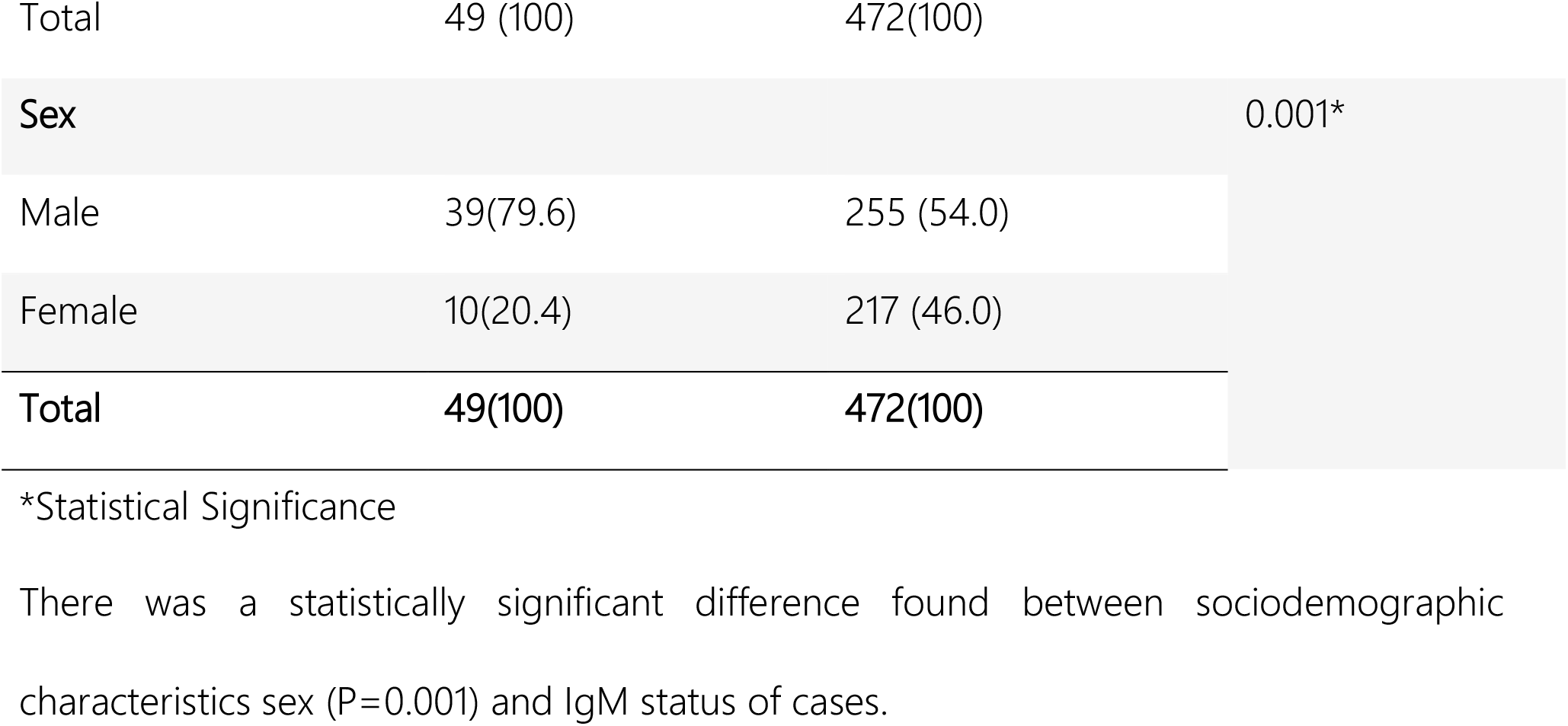
Association between sociodemographic characteristics and IgM status

| Characteristics | IgM Status |  |  |  | P value |
| --- | --- | --- | --- | --- | --- |
|  | Positive (%) | Frequency | Negative (%) | Frequency |  |
| Age |  |  |  |  | 0.957 |
| <30 | 20 (40.8) |  | 138 (40.0) |  |  |
| 30-49 | 18 (36.7) |  | 135 (38.8) |  |  |
| >49 | 11 (22.4) |  | 81 (21.2%) |  |  |

|  |  |  |  |
| --- | --- | --- | --- |
| Total | 49 (100) | 472(100) |  |
| **Sex** |  |  | 0.001* |
| Male | 39(79.6) | 255 (54.0) |  |
| Female | 10(20.4) | 217 (46.0) |  |
| **Total** | **49(100)** | **472(100)** |  |

\*Statistical Significance
There was a statistically significant difference found between sociodemographic characteristics sex (P=0.001) and IgM status of cases.

**Table 5.**
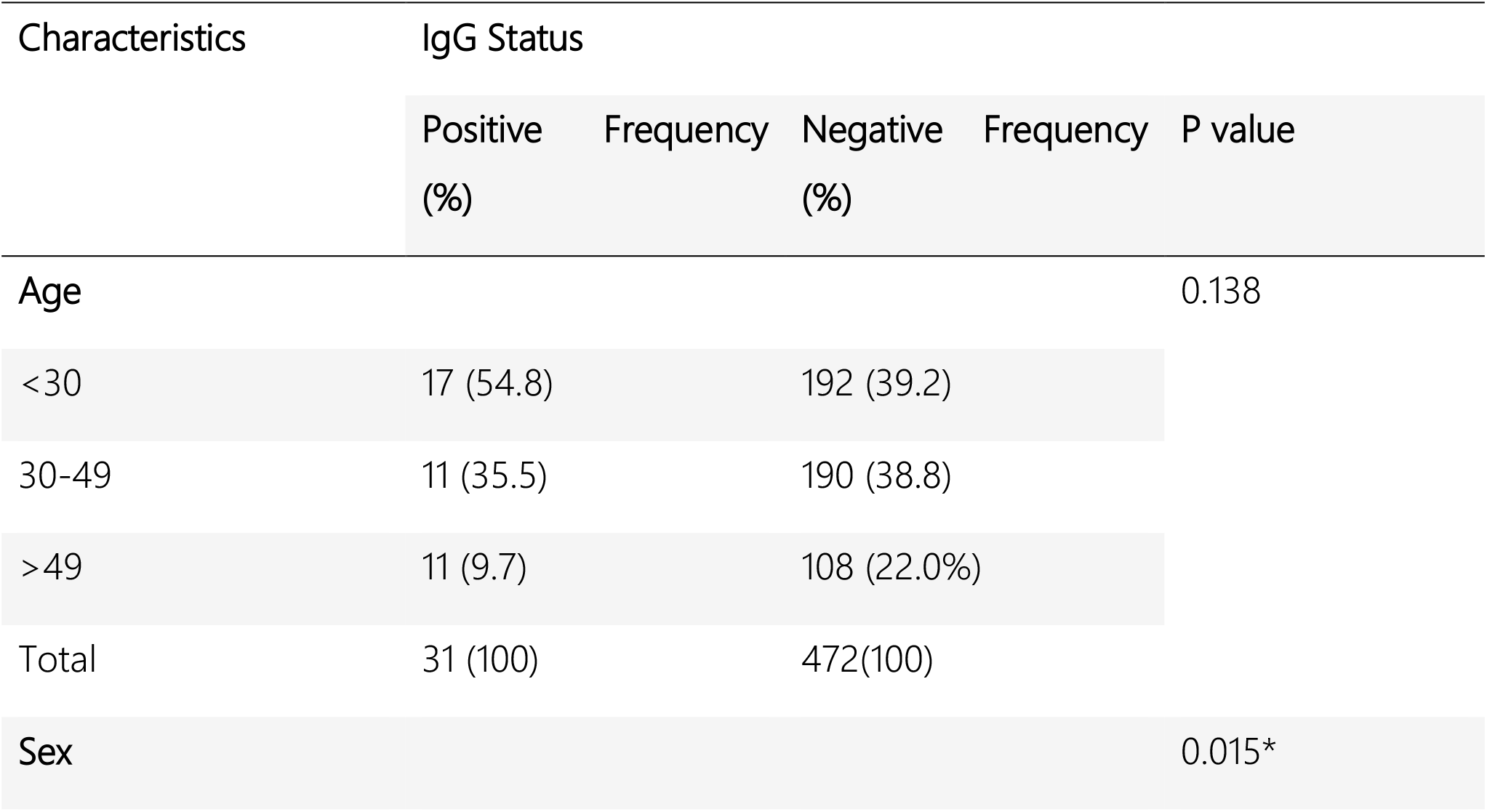
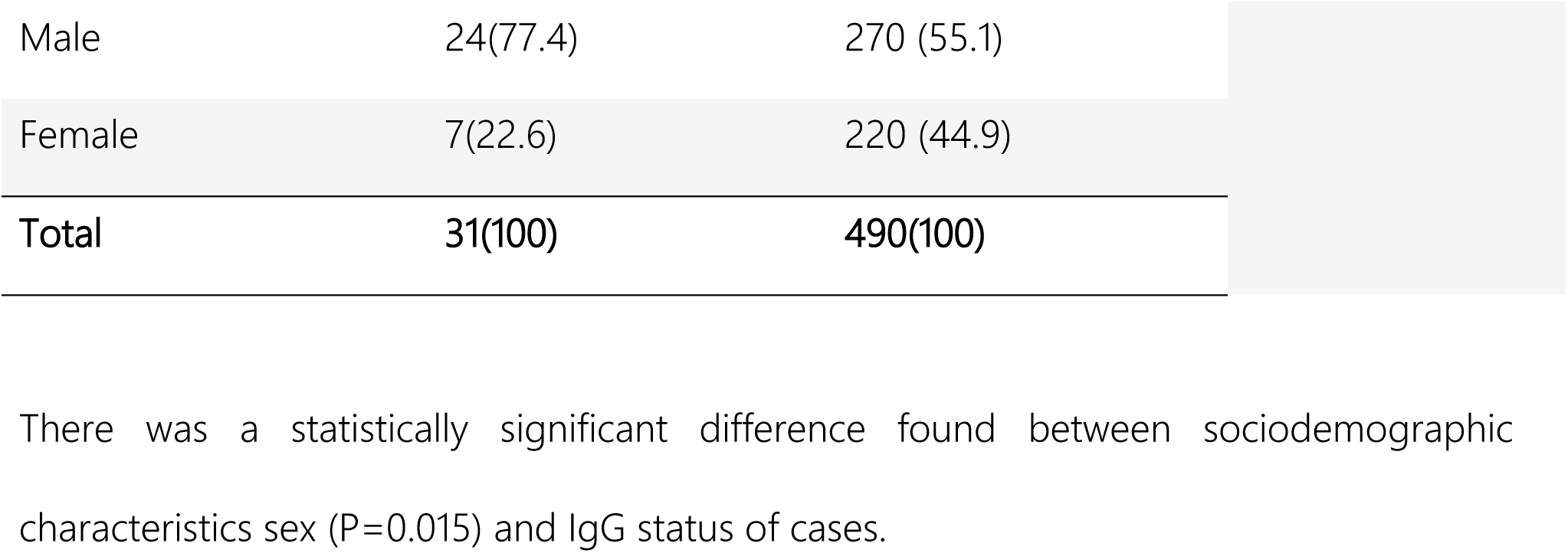
Association between sociodemographic characteristics and IgG status

| Characteristics | IgG Status |  |  |  | P value |
| --- | --- | --- | --- | --- | --- |
|  | Positive (%) | Frequency | Negative (%) | Frequency |  |
| **Age** |  |  |  |  | 0.138 |
| <30 | 17 (54.8) |  | 192 (39.2) |  |  |
| 30-49 | 11 (35.5) |  | 190 (38.8) |  |  |
| >49 | 11 (9.7) |  | 108 (22.0%) |  |  |
| Total | 31 (100) |  | 472(100) |  |  |
| **Sex** |  |  |  |  | 0.015* |

|  |  |  |
| --- | --- | --- |
| Male | 24(77.4) | 270 (55.1) |
| Female | 7(22.6) | 220 (44.9) |
| **Total** | **31(100)** | **490(100)** |

There was a statistically significant difference found between sociodemographic characteristics sex (P=0.015) and IgG status of cases.

**Table 6.**
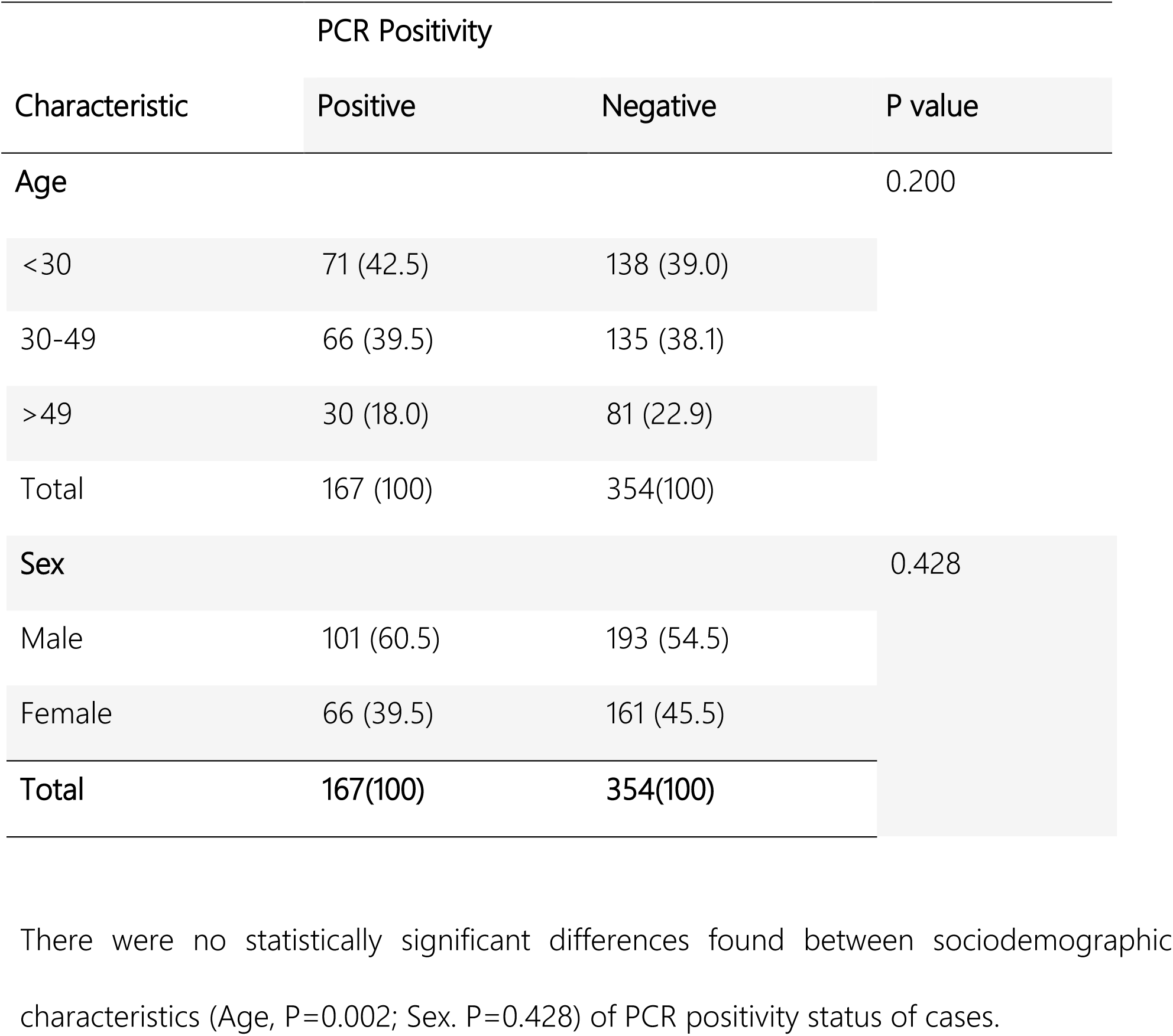
Association between sociodemographic characteristics and PCR positivity

### IgM and IgG reactivities in PCR-confirmed COVID-19 patients

Altogether 24 of 166 (14.5%) samples from PCR confirmed COVID-19 patients tested IgM positive and 26 tested (15.7%) IgG positive (Tables 2 and 3).

### Assay sensitivity, specificity and accuracy

Based on the results described above and summarized in Tables 2 and 3, the assay showed a sensitivity of 14.4% (24/167) and 15.6% (26/167) for IgM and IgG, respectively. The assay showed an overall specificity of 92.9% (329/354, 25 false positive) and 98.6% (349/354, 5 false positive) for IgM and IgG, respectively. Using PCR-positive cases as true positives, the accuracy of the test was 67.8% (353/521) and 72.0% (375/521) for IgM and IgG, respectively. The positive and negative predictive values (the likelihood of being a case given a positive test result, and the likelihood of being healthy given a negative test result) for IgM were 48.9% (24/49) and 69.7% (329/472), respectively. For IgG, the corresponding values were 83.9% (26/ 31) and 71.2% (349/490).

## Discussions

The present study evaluated a commercial rapid test for detection of SARS-CoV-2-specific IgM and IgG. For the evaluation, samples from COVID-19 cases were obtained during disease condition and previously confirmed by PCR, were used as the true positives. According to the manufacturer of the PCR test, the specificity had been evaluated and was found to be 100% (95% CI:94.20% to 100%) for both IgM and IgG, while the sensitivity evaluated on COVID-19 cases was 87.3% (95% CI: 80.40% to 92.0%) for both IgM and IgG. Our results showed an overall very low sensitivity of IgM (14.4%) and IgG (15.6%). A study carried out by Li et al. in China in contrast to our study findings reported an overall high testing sensitivity of 88.7% and 90.6% specificity. The specificity results for our study for IgM (92.9%) and IgG (98.6%) were higher than what was reported in Li *et al* study.

There were 25 false IgM positive samples and 5 false positive IgG results observed from the healthy participants. This indicates that a cross-reaction to another coronavirus might have taken place. Previous literature has documented the occurrence of serological cross-reactions between SARS-CoV and SARS-CoV-2 (Wan WY 2020). Globally there are some other types of coronaviruses that affect humans that are endemic or even occur as epidemics. These include the 229E, HKU1, NL63 and OC43 strains. The picture of significant false positivises observed in this study may possibly be due the cross reaction from a previous infection with one of the strains of those other viruses mentioned above. It cannot be said for certain the commonality of the corona viruses as the agents responsible for the flu, but it has been estimated that about 20% of cases of the flu could be as a result of the coronaviruses (Bende M, 1989). The specificity for IgG detection of the rapid test evaluated in this study was similar to what was reported in another study of an enzyme-linked immunosorbent assay (ELISA), which had a specificity 97.5% (Zhao R 2020). However, the sensitivity of our study was very low as compared with that of the ELISA which was also 97.5%. This study generally showed a poor sensitivity performance of antibody tests for both IgM and IgG. The negative predictive values for the rapid tests in this study were not very high, indicating that this rapid test would not be useful for detecting past infections and possible immunity. It is not known when infection occurred for individuals in this study. The timing of the development of SARS-CoV-2–specific antibodies is variable. The humoral response kinetics to SARS-CoV-2 infection have not been fully understood, however it has been shown that reactive IgA, IgM, and IgG antibodies have been detected as soon as 1 day after symptom onset (Guo, 2020). In other previous studies, the antibodies were detected 10 to 15 days after symptom onset. The median time to the development of total antibody, IgM, and IgG has been estimated as 11, 12, and 14 days, respectively (Zhao,2020; Wu F,2020). We also investigated the possible associations between IgM, IgG and PCR positivity and sociodemographic characteristics. The associations found were those between IgM and IgG status and gender. According to the previous studies, gender has not been shown to have significant association with antibodies to SARSCoV-2. A study conducted in Spain did not show any statistically significant association with presence of IgM and IgG antibodies to SARS-CoV-2 and gender (Garcia-Basteiro 2020). Another study conducted in the United States of America similarly did not find any association between the presence of IgM and IgG antibodies to SARS-CoV-2 and gender (Havers FP, 2020). In another study conducted by Zeng *et al*. among 331 patients confirmed SARS-CoV-2 infection, it was found that level of IgG antibody in mild, general and recovering patients did not show any difference between males and females. However, the average IgG antibody level in female patients tended to be higher than that of in male patients in severe infections. It was observed that in comparison to male patients, most of the female patients generated a relatively high level of SARS-CoV-2 IgG antibody in severe infection. It was also reported that, the generation of IgG antibody in the females tended to be stronger than the male patients in the early phase of the disease. There are inconsistent findings from various studies on the associations between gender and SARS-CoV-2 antibody positivity. It is necessary for additional studies to be carried out to explore further and validate the relationship between gender and SARS-CoV-2 antibody positivity.

## Recommendation and Conclusion

Although rapid antibody test kit has been found to be simple, inexpensive and faster in assessing COVID-19 infection in Humans, the specificity and sensitivity of the Innovita test kit for detecting SARS-CoV2 antibody as claimed by the manufacturer need to be reevaluated. The Rt-PCR still remain the gold standard as recommended by WHO for the diagnosis of SARS CoV2 infection until a more specific, sensitive and reliable rapid test kit has been developed and recommended by WHO for use at point of entry, point of care and for general community mass screening program for COVID-19 antibody.

## Data Availability

All relevant data are included in the manuscript as required.

## Acknowledgement

The authors acknowledge the Ministry of Health (MOH) Kaduna State for the approval of this pilot study, Barau Dikko Teaching Hospital (BDTH) and the Laboratory Pillar Kaduna State COVID-19 Rapid Response Team for their assistance and expertise

## APPENDIX I

### PARTICIPATION INFORMATION LEAFLET

Serial no………………

### Investigators

**Study Title**: Validation of SARS-CoV2 Antibody Tests

### Introduction

The emergence of the Corona virus disease 2019 (COVID-19) pandemic has had significant impact on global health indices. Understanding the underlying mechanism(s) of the disease and the full clinical picture are still ongoing.

The gold standard for diagnosis of COVID-19 is Real Time Polymerase Chain Reaction (RT-PCR) which identifies the virus. There are new test kits that are commercially available that attempt to identify if one has been previously infected by identifying special proteins called antibodies formed by the body in response to infection. The ability of these tests to accurately identify persons infected with COVID-19 is still being evaluated.

This study seeks to compare the accuracy of a test kit (INNOVITA) against the gold standard for screening and diagnosis

The findings from this study will hopefully make COVID-19 testing cheaper and less associated with stigma

Your participation includes sample collection (4ml of venous blood). Needle pricks are associated with temporary pain and every effort will be made to minimize this. Tests performed will be at no cost to you. Your participation is entirely voluntary and you may withdraw from the study at any time without bearing consequences. The ethical committee of Kaduna State Ministry of Health has given approval for this study to be carried out. If you are willing to participate in this study, please complete the consent form attached.

## APPENDIX II

### CONSENT FORM FOR RESEARCH PARTICIPANTS

I…………………………………………of………………………………………… (address) agree to participate in the study on validation of SARS-CoV2 Antibody Tests.

The full procedure for the tests has been explained to me and I understand that my blood sample and oral/nasal swab will be taken.

I make this consent without being subjected to any pressure. Participant’s name & signature………………………………Date…………………………

Witness’name & signature……………………………………Date…………………………

Principal Investigator’s name & signature………………………………Date…………………………

Contact Phone no:

## APPENDIX III

RT-PCR test result …………………

Date of test (DD/MM/YYYY) …………

Interval from onset of symptoms (fever, cough, shortness of breath etc) (in days) ……………

Antibody test result

SARS-CoV2 Antibody Result: IgG [ ]

IgM [ ]

